# Selective Molecular and Network Architecture Features Underlie Brain Cortical Atrophy in Dementia with Lewy Bodies

**DOI:** 10.64898/2026.05.26.26354105

**Authors:** Aline Delva, Stephen Joza, Christina Tremblay, Andrew Vo, Marie Filiatrault, Madeleine Carrier, John-Paul Taylor, John T. O’Brien, Michael Firbank, Alan Thomas, Paul C. Donaghy, Richard Camicioli, Howard Chertkow, Alain Dagher, Ronald B. Postuma, Shady Rahayel

## Abstract

**BACKGROUND:** Dementia with Lewy bodies shares clinical and pathological features with both Parkinson’s disease and Alzheimer’s disease, but the local biological factors that render specific cortical regions vulnerable to atrophy remain poorly defined. In particular, it is unclear whether cortical thinning in dementia with Lewy bodies reflects generic neurodegenerative mechanisms, processes shared with Parkinson’s disease and Alzheimer’s disease, or dementia with Lewy bodies-specific molecular and network susceptibilities.

**METHODS:** A total of 89 patients with dementia with Lewy bodies and 89 matched controls underwent T1-weighted brain MRI. Scans were processed to generate surface-based cortical thickness maps. Regional cortical thickness estimates, after slice-by-slice manual correction, were mapped to gene expression data from healthy postmortem human brains to identify transcriptomic signatures associated with decreased thickness in dementia with Lewy bodies. We assessed whether genes whose expression was increased with regional thinning converged onto established Parkinson’s disease- and Alzheimer’s disease-related pathways and isolated genes uniquely implicated in dementia with Lewy bodies. Spatial annotation mapping was then used to test whether patterns of cortical thinning overlapped with in vivo neurotransmitter system distributions and whether the observed thickness pattern was constrained by large-scale structural connectivity, consistent with a network-based propagation process.

**RESULTS:** Cortical thinning predominated in regions that, in the healthy brain, show higher expression of genes involved in mitochondrial function and synaptic transmission. The transcriptomic profile associated with thinning significantly overlapped with genes belonging to Parkinson’s disease and Alzheimer’s disease pathways, supporting shared pathogenic mechanisms across Lewy body and Alzheimer-type neurodegeneration. However, 90 genes associated with cortical thinning did not overlap with Parkinson’s disease or Alzheimer’s disease pathways and were enriched for GABAergic signalling. Spatial mapping analyses showed that regions with greatest thickness reductions colocalized with GABA_A_, serotoninergic 5-HT_1A_, 5-HT_1B_, 5-HT4, and dopaminergic D2 receptor distributions, and that the thickness pattern followed structural connectivity.

**CONCLUSIONS:** MRI-derived cortical thickness changes in dementia with Lewy bodies reflect selective molecular and network vulnerabilities rather than a non-specific degenerative process. Mitochondrial and synaptic genes, together with a distinct GABAergic association and connectivity constraints, delineate mechanisms explaining why some cortical territories are more affected in dementia with Lewy bodies.

## Background

Dementia with Lewy bodies (DLB) is a common form of late life neurodegenerative dementia,^1,2^ characterized by parkinsonian features, visual hallucinations, cognitive fluctuations, and dream enactment behaviours, along with dementia.^1, 2^ As synucleinopathy, DLB involves progressive accumulation of α-synuclein aggregates (Lewy bodies and neurites) mainly within the cerebral cortex and limbic system.^2, 3^ Co-occurring β-amyloid pathology is common^2, 4, 5^ and has been associated with worse prognosis and accelerated brain atrophy.^6-8^ Structural MRI studies have consistently shown widespread cortical atrophy in DLB compared to healthy ageing,^9, 10^ typically posterior including the parietal, occipitotemporal, and posterior cingulate cortices.^10^ This pattern partially resembles that of Alzheimer’s disease (AD), though it tends to be less diffuse and often spares medial temporal lobes.^9^ These structural changes correlate with cognitive decline in DLB,^9, 10^ yet the local biological factors that drive selective vulnerability to atrophy remain poorly understood.

Emerging evidence suggests that neurodegeneration is not random but shaped by the brain’s intrinsic biological architecture, including regional gene expression patterns and network connectivity.^11, 12^ These intrinsic features may predispose certain areas to pathology and make them more susceptible to neurodegeneration.^13-16^ In animal models, misfolded alpha-synuclein spread occurs via anatomically connected circuits, supporting the concept of prion-like propagation.^13-16^ This spread happens within a landscape of selective vulnerability, where regions differ in susceptibility based on their molecular profiles.^13, 17, 18^ In Parkinson’s disease (PD) and idiopathic/isolated REM sleep behaviour disorder (iRBD),^19^ a sleep disorder where over 90% of individuals eventually develop a manifest synucleinopathy including DLB,^19^ imaging transcriptomics and network propagation models have shown that MRI-derived cortical thickness reductions preferentially affect areas with high expression of mitochondrial, synaptic, or autophagy-related genes, as well as those embedded in densely connected brain networks.^20-23^ In multiple system atrophy, atrophied regions are enriched for oligodendrocyte-related genes,^24^ while in Alzheimer’s disease, affected regions show enrichment for protein remodelling pathways, with *APOE* emerging as a key gene.^22^ These findings demonstrate how imaging-genomic approaches can identify disease-specific mechanisms of neurodegeneration.

DLB overlaps with both PD and AD in terms of clinical features, risk genes, pathology, and MRI-based measures of neurodegeneration.^1, 9, 25, 26^ Motor features and synuclein aggregation are similar between DLB to PD, while cognitive decline and amyloid deposition are more similar to AD.^1, 4, 27, 28^ The DLB atrophy pattern also partially overlaps with PD and AD.^1, 9, 28^ Moreover, a structural MRI-derived atrophy signature has been shown to predict DLB in individuals with iRBD, underscoring the potential of brain atrophy patterns as prognostic biomarkers.^29^ However, no study has yet examined DLB-related atrophy in the context of the brain’s molecular and network architecture. Increasingly, rich multimodal datasets from well-characterized healthy individuals provide a valuable normative reference for contextualizing disease-specific brain changes. Leveraging such normative data allows us to identify whether patterns of atrophy in DLB align with specific molecular vulnerability or network organization in the healthy brain. This approach could help assess whether the observed atrophy reflects biological mechanisms shared with PD and AD, or whether distinct features underlie DLB-specific vulnerability. Addressing this question may clarify which susceptibility features underly DLB-related neurodegeneration and inform the development of targeted therapeutic strategies.

In this study, we aim to investigate the normative biological factors underlying regional vulnerability to cortical atrophy in DLB. We first quantified cortical thickness, surface area, and volume patterns in a multicentre DLB cohort compared to matched controls. We then applied imaging transcriptomics to assess whether abnormal regions were associated with the brain’s spatial gene expression pattern and enriched for genes implicated in PD and AD pathways. We next assessed the enrichment patterns of genes that were non-overlapping with PD and AD pathways to identify processes that may be more uniquely implicated in DLB-associated neurodegeneration. Spatial annotation mapping was applied to investigate whether cortical changes overlapped with specific neurotransmitter systems distributions using a large collection of PET-based tracer maps. Finally, we investigated whether the DLB pattern of cortical changes was constrained by structural and functional brain connectivity. We hypothesized that the healthy transcriptomic patterns overexpressed in brain regions vulnerable to neurodegeneration in DLB would partly overlap with those implicated in both PD and AD, with enrichment of mitochondrial and synaptic processes. Consistent with this framework, we further expected neurodegeneration-prone regions in DLB to occupy highly connected white matter networks and to show preferential enrichment for specific neurotransmitter systems.

## Methods

Figure 1 summarizes the main processing and analyses. We first mapped the cortical thickness, surface, and volume patterns in DLB using structural MRI (Fig.1a). We then applied imaging transcriptomics to identify biological mechanisms associated with the cortical patterns (Fig.1b). Next, we assessed spatial correspondence between the patterns in DLB and brain maps of neurotransmitter systems (Fig.1c) to investigate potential involvement of specific neurotransmitters. Finally, connectomic analyses determined whether structural and functional connectivity constrained the spatial distribution of DLB cortical patterns (Fig.1d).

**Figure 1.**
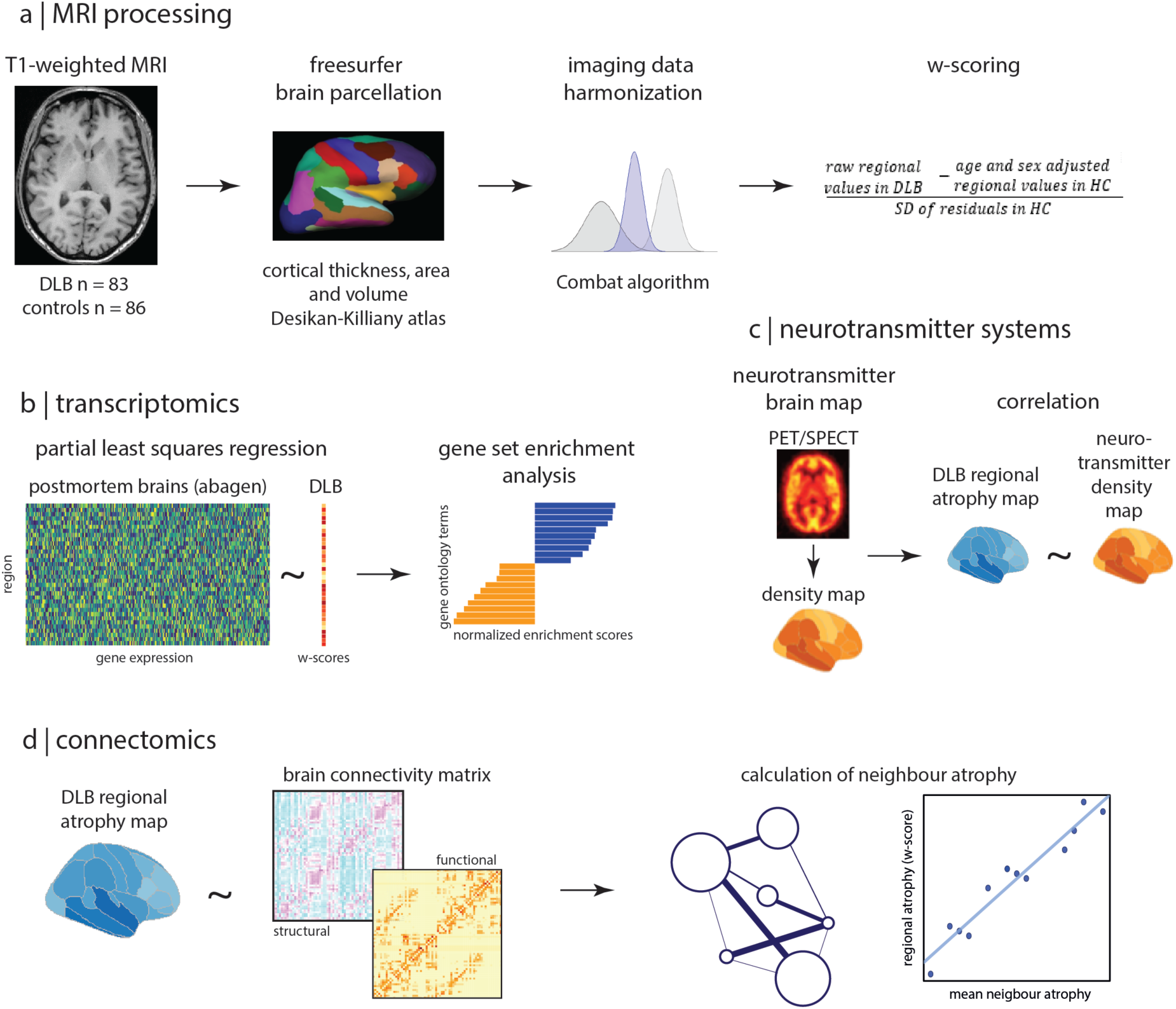
Overview of the pipeline. (a) T1-weighted MRI scans from patients with DLB and healthy controls were processed. Surface maps were parcellated into the Desikan-Killiany atlas to calculate regional cortical thickness, volume, and surface area. Scan harmonization was used to correct for inter-scanner variability. Regional W-scores in DLB were calculated by subtracting age- and sex-adjusted healthy control regional values, dividing by the SD of control residuals. (b) For imaging transcriptomics analysis, regional gene profiles were extracted from postmortem brain data using abagen. Partial least squares regression was then used to quantify the association between the average DLB cortical vector and regional gene expression. Gene set enrichment analysis was done to identify significant gene ontology terms enriched in relation to regional brain changes in DLB. (c) Previous PET imaging data were used to calculate different neurotransmitter system density maps. These density maps were correlated with regional cortical alterations in DLB. (d) The effect of structural and functional brain connectomics on cortical changes in DLB was assessed by correlating regional cortical change in DLB and average change in structurally and functionally connected neighbors, weighted by the respective connectivity matrices.

### Participants

A total of 178 participants (89 with DLB and 89 age- and sex-matched healthy controls) were included from two datasets: 132 (75 with DLB) from the University of Newcastle (United Kingdom) as part of three longitudinal studies: TMSS, CATFIELD, and AMPLE cohorts. The additional 46 participants (14 with DLB) were part of the Comprehensive Assessment of Neurodegeneration and Dementia (COMPASS-ND) study, a Canadian multicentre, prospective initiative of the Canadian Consortium on Neurodegeneration in Aging (CCNA). All DLB participants met diagnostic criteria for probable DLB,^1^ based on detailed neurological and neuropsychological assessments. Core clinical features (i.e. parkinsonism, visual hallucinations, cognitive fluctuations, and iRBD) were systematically assessed in both cohorts (Supplementary Table 1 for cohort details). Global cognition was evaluated using the Mini-Mental State Examination (MMSE) in the Newcastle cohort and the Montreal Cognitive Assessment (MoCA) in the CCNA cohort. Parkinsonian motor features were assessed using the Unified Parkinson’s Disease Rating Scale part III (UPDRS-III) in Newcastle and the Movement Disorder Society (MDS)-revised UPDRS-III in the CCNA cohort. Healthy controls were community-dwelling individuals without sign of cognitive decline or neurodegenerative diseases. Demographic and clinical characteristics were compared between groups using Student’s t-tests for continuous variables and χ² tests for categorical variables. MoCA and MMSE scores, as well as UPDRS-III and MDS-UPDRS-III scores, from the different cohorts were harmonized using validated conversion algorithms to enable cross-cohort comparisons.^36^ In the DLB group, associations between regional cortical changes and clinical measures (global cognition and parkinsonian motor features) were assessed using ANOVAs adjusted for age and sex, with FDR correction for multiple comparisons.

All participants provided written informed consent in accordance with institutional ethics protocols. The original study protocols received approval from local research ethics committees, and the present study was approved by the Research Ethics Boards of the McGill University Health Centre and the Quebec Integrated University Centre for Health and Social Services of Northern Island of Montrfeal.

### MRI

#### MRI acquisition and processing

T1-weighted MRI scans were acquired at each centre (see Supplementary Materials for details). Detailed processing steps are available in the Supplementary materials. In short, T1-weighted MRI scans were processed using FreeSurfer v7.1.1 to generate vertex-wise maps of cortical thickness, surface area, and cortical volume, following following procedures from previous studies such as skull stripping, surface reconstruction, anatomical parcellation, and intensity normalization. All reconstructions were visually inspected and manually corrected slice by slice by trained raters (A.De., S.J., S.R.). Maps with major segmentation errors (rating > 2 on a 4-point scale) were excluded. Of the 178 available scans (89 DLB, 89 age- and sex-matched controls), 9 (5%) were excluded due to failed FreeSurfer processing or quality control (6 DLB, 3 controls).

To assess morphological differences between DLB patients and controls, we conducted region-and vertex-wise analyses of cortical thickness, surface area, and cortical volume separately. Surface-based analyses were performed to provide finer spatial resolution compared to region-wise analyses. Cortical thickness maps were smoothed using a 15-mm full-width at half-maximum (FWHM) Gaussian kernel, and a general linear model (GLM) was fitted at each cortical vertex to test for group differences while controlling for age and site. Sex was not in the GLM due to class imbalance, which would give unstable estimates. However, the sex effect was controlled for region-based analyses using W-scoring based on control data. Statistical significance was assessed using cluster-wise Monte Carlo simulation, with the cluster-wise threshold set at *P*_cluster_ < 0.05 and the vertex-wise cluster threshold set at *P*_vertex_ < 0.001.^30^

For the complementary region-wise analysis, cortical surface maps were smoothed with a 15-mm full-width at half-maximum (FWHM) Gaussian kernel and parcellated into 68 regions (34 per hemisphere) using the Desikan-Killiany atlas.^31^ To account for scanner-related variability, morphometric measures were harmonized using the ComBAT algorithm,^32^ a batch-effect correction method validated for multi-site MRI datasets.^20, 33^ ComBAT was applied with age, sex, and diagnosis as covariates to preserve biological variability. Group differences in ComBAT-corrected regional morphometry were tested using analysis of variance (ANOVA), controlling for age and sex across all models. Age and sex effects in normal aging were regressed out using W-score modelling based on matched controls.^34, 35^ Negative W-scores indicated decreased measurement compared to controls (atrophy for thickness and volume), while positive scores reflected enlargement compared with controls. The regional W-scores were used in the imaging transcriptomics, cortical thickness, surface area, and volume analyses. Estimated total intracranial volume (eTIV) was included as covariate in all models assessing surface area and cortical volume. ANOVAs were conducted separately for each region, and significance was determined using FDR correction across comparisons (P_FDR_ < 0.05, two-tailed).

#### Imaging transcriptomics

Imaging transcriptomics was used to associate neuroimaging patterns with normative gene expression profiles derived from post-mortem human brain tissue.^36-38^ Notably, we applied this approach to test whether cortical thickness, surface area, and volume in DLB overlapped with spatial patterns of gene expression in the healthy brain, using 68 cortical regions from the Desikan-Killiany atlas.

We used gene expression data from the Allen Human Brain Atlas, which provides microarray-based profiles for over 20,000 genes across 3,702 brain tissue samples from six neurotypical adult donors.^39, 40^ Data were processed with the *abagen* toolbox (v0.1.3).^39^ Probes were reannotated^36^ and filtered to exclude those with expression levels below background in ≥50% of samples.^41^ For genes with multiple probes, the one with the highest differential stability, defined as the average Spearman correlation of expression across donors, was retained.^42^ MNI coordinates of tissue samples were updated via non-linear registration using Advanced Normalization Tools. After probe filtering, reannotation, and normalization, expression data for 15,633 genes were mapped to the same 68 cortical regions used in the MRI analyses; each sample was assigned to a Desikan-Killiany region if its coordinates were within 2 mm of a given parcel,^31^ constrained by hemisphere and gross anatomical division to minimize misclassification.^36^ Unassigned samples were excluded. Expression values were normalized across genes within each sample using a robust sigmoid function, rescaled to the unit interval, and then normalized again across samples. Regional gene expression values were averaged first across samples within each region per donor, then across donors. Due to limited right hemisphere sampling (available in only two donors), left hemisphere values were mirrored to the right.

#### Partial least squares regression

Partial least squares regression was used to identify latent variables (LVs) that capture maximal covariance between regional gene expression and cortical thickness, surface area, or volume in DLB, treated separately.^43^ Specifically, we tested whether LVs derived from the gene expression matrix **X** (68 cortical regions x 15,633 genes) explained significant variance in the cortical vector **Y** (W-scored cortical values averaged across participants for the same 68 regions). To assess statistical significance, we generated 10,000 permutations comparing the variance explained by each LV against two null models: 1) a random null, created by shuffling cortical values across regions, and 2) a spatial null, generated via spherical reassignment of cortical values to preserve spatial autocorrelation (i.e., accounting for the similarity between values in adjacent regions).^44^ An LV was considered significant if it explained more variance than 95% of both null distributions.

To assess the biological relevance of gene expression patterns associated with cortical thickness, surface area, or volume in DLB, we applied a bootstrapping approach. The gene expression matrix **X** and cortical vector **Y** were randomly shuffled 5,000 times to generate null distributions for each LV. For each significant LV, we computed a bootstrap ratio for every gene, defined as the ratio of its expression weight to its standard error. These ratios, interpreted as z-scores, were sorted from highest (genes overexpressed in regions with less changes) to lowest (genes overexpressed in regions with the most changes such as atrophy) and used as input for gene enrichment analysis.

#### Gene enrichment analyses

Gene set enrichment analysis was performed to identify biological processes and cellular components enriched in the ranked gene lists.^45^ Enrichment was conducted using WebGestalt 2024^46, 47^ with the Gene Ontology (GO) database (October 2023 release, containing 45,003 terms). Analyses were limited to GO categories containing between 5 and 2,000 genes and 10,000 permutations were used to calculate false discovery rate (FDR)-corrected P-values. For each significant GO term, we extracted the normalized enrichment score, gene set size, and number of leading-edge genes. Negative normalized enrichment scores indicated enrichment among genes overexpressed in atrophied regions, while positive scores indicated enrichment in regions with relative preservation. To ensure robustness, enrichment results were independently replicated using the PANTHER 18.0 classification system.^48^

To test overlap with Lewy body dementia genetics, we compiled the previously identified 69 Lewy body dementia risk genes from a published gene-level GWAS.^49^ Gene symbols were harmonized to our Allen Human Brain Atlas-based transcriptomic matrix, and gene weights relating regional expression to the DLB cortical map were obtained with the bootstrapping framework described above. We classified genes as overexpressed in atrophied regions if their bootstrap ratio was below -1.96, corresponding to being outside the central 95% of the standard normal distribution (two-tailed *P* = 0.05).

### Pathway-specific enrichment of DLB-related cortical changes

To determine whether cortical changes in DLB are driven by mechanisms also involved in PD and AD, we performed a pathway enrichment analysis on the ranked gene list derived from the imaging transcriptomics analysis. We used the Kyoto Encyclopaedia of Genes and Genomes (KEGG) pathways, which comprise curated gene sets representing well-defined biological processes, including those implicated in neurodegenerative diseases.^50^ We tested whether genes associated with DLB-related cortical changes in our study (ranked by bootstrap ratio weights from LV1) were overrepresented in certain pathways, particularly PD- and AD-related KEGG pathways. We focused on negatively weighted genes, as these are more highly expressed in regions showing cortical changes in DLB.

Since KEGG pathways often share overlapping genes, we next performed a redundancy analysis across all significantly enriched pathways. Genes were grouped into four categories: 1) unique to PD-related pathways, 2) unique to AD-related pathways, 3) shared between PD and AD, and 4) not part of either (representing potential DLB-specific processes). To explore the functional relevance of each group, we conducted GO biological process enrichment in each group against the full gene set as background.

To assess the brain relevance of the DLB-specific genes (i.e., those linked to cortical changes but absent from PD or AD pathways), we further investigated their tissue-specific expression using RNA sequencing data from the Genotype-Tissue Expression (GTEx) Project (release V10).^51^ This dataset includes transcriptomic profiles across 68 tissue types from nearly 1,000 healthy post-mortem donors. For each gene, median expression (in transcripts per million, TPM) was computed across brain/CNS tissues (N=13) and compared to non-brain tissues (N=55) using a Wilcoxon rank-sum test (Supplementary Table 2 for list of tissues). We derived mean TPM values, log_2_ fold-change (brain vs. non-brain), raw P-values, and FDR-adjusted q-values. Genes with q < 0.05 were considered significantly enriched in the brain/CNS tissue. To characterize the functional roles of these brain-enriched DLB-specific genes, we repeated GO enrichment analysis in these genes against all genes as background.

### Structural and functional connectivity analysis

To evaluate whether connectivity determined cortical changes in DLB, we assessed whether structural and functional connectivity constrained regional patterns of cortical thinning. Specifically, we tested whether the cortical thickness W-scores of a given region (‘node’) was associated with the average W-score of its connected neighbour nodes, based on normative connectome data. Structural and functional connectivity matrices, using the 68 cortical regions from the Desikan-Killiany atlas as nodes, were derived from an independent sample of healthy participants who underwent cutting-edge high-resolution diffusion-weighted imaging and resting-state functional MRI as part of the Human Connectome Project.^52^

For each region *i*, we computed a connectivity-weighted average cortical score across its neighbours (*T_i_*), using either structural or functional connectivity as weights. Structural neighbourhood cortical change was defined as previously described ^53^:

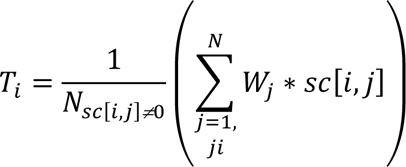

where *N*_*sc*[*i,j*]≠0_ is the total number of non-zero weighted connections between regions, *W_j_* is the cortical thickness W-score of neighbour *j*, and *sc[i,j]* is the strength of the structural connection between regions *i* and *j*. Similarly, functional neighbourhood cortical change was defined as:

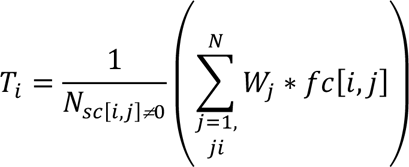

where the functional connectivity strength *fc[i,j]* was used to weight the cortical thickness of regions structurally connected (sc=0) to region *i*. This evaluated whether functional coupling among anatomically connected regions modulated the expression of cortical thinning in DLB.

We then calculated Spearman’s correlation between each region’s W-score and its connectivity-weighted neighbourhood cortical change *T_i_*, separately for structural and functional connectivity. Statistical significance was evaluated using 10,000 spin permutations to generate spatially constrained null distributions, implemented via the *netneurotools* toolbox.

### Mapping DLB cortical thickness pattern to neurotransmitter systems

To assess whether DLB-related cortical thinning might be related with different neurochemical systems, we tested whether the spatial pattern of cortical change corresponded to regional distributions of neurotransmitter receptors, transporters, and synaptic density in the healthy brain. We used the *neuromaps* toolbox^54^ to obtain PET- and SPECT-derived maps for 9 neurotransmitter systems and one synaptic marker, parcellated into the 68 cortical regions of the Desikan-Killiany atlas. These included GABAergic (GABA_A_), glutamatergic (mGluR5), cholinergic (α4β2, M1, vesicular acetylcholine transporter [VAChT]), dopaminergic (D1, D2, dopamine transporter [DAT]), noradrenergic (noradrenaline transporter [NET]), serotoninergic (5-HT_1A_, 5-HT_1B_, 5-HT_2A_, 5-HT_4_, 5-HT_6_, serotonin transporter [5-HTT]), endocannabinoid (CB_1_), opioid (µ), histaminergic (H_3_), and synaptic density (SV2A) maps.^54^ Full methodological details for each tracer are provided in Supplementary Table 3.^54^ When multiple maps were available for the same tracer, we computed a weighted average based on sample size. All maps were z-scored across regions to standardize values. We then computed Spearman’s correlations between regional cortical thickness W-scores in DLB and each neurochemical map. Resulting p-values were corrected for multiple comparisons using FDR. Significant correlations were further tested against 10,000 random and spatial distributions using *netneurotools* (https://github.com/netneurolab/netneurotools).

### Availability of data and materials

The dataset supporting the conclusions of this article were obtained from multiple sources. Access to the Newcastle cohort is governed by institutional policies and requires approval from the study investigators. For the CCNA dataset, qualified researchers may request access to COMPASS-ND data through the CCNA. Group-average parcellated maps for DLB and control participants can be shared upon reasonable request. All software tools used in the analyses are publicly available and referenced in the Methods section.

## Results

### Participants

A final sample of 169 participants comprising 83 DLB patients (mean age: 76.8 ± 6.4 years; 24 females) and 86 controls (75.0 ± 6.3 years; 33 females) was included in our analyses. There were no significant group differences in age (P = 0.06) or sex distribution (P = 0.26). As expected, DLB patients showed greater cognitive and motor impairment compared to controls. MoCA and MMSE scores were significantly lower in the DLB group (P < 0.0001) and UPDRS-III scores were higher (DLB: 33.0 ± 18.5 vs. Controls: 2.7 ± 3.0, Newcastle cohort, P < 0.0001).

In the CCNA cohort, the mean MDS-UPDRS-III score for DLB patients was 55.7 ± 22.2 (control data unavailable). Core clinical features were prevalent among DLB patients: 67% reported visual hallucinations, 73% cognitive fluctuations, 77% parkinsonism, and 52% had a history suggestive of iRBD (all *P* < 0.0001 vs. controls). Full demographic and clinical details are provided in Table 1 and Supplementary Table 4.

**Table 1.**
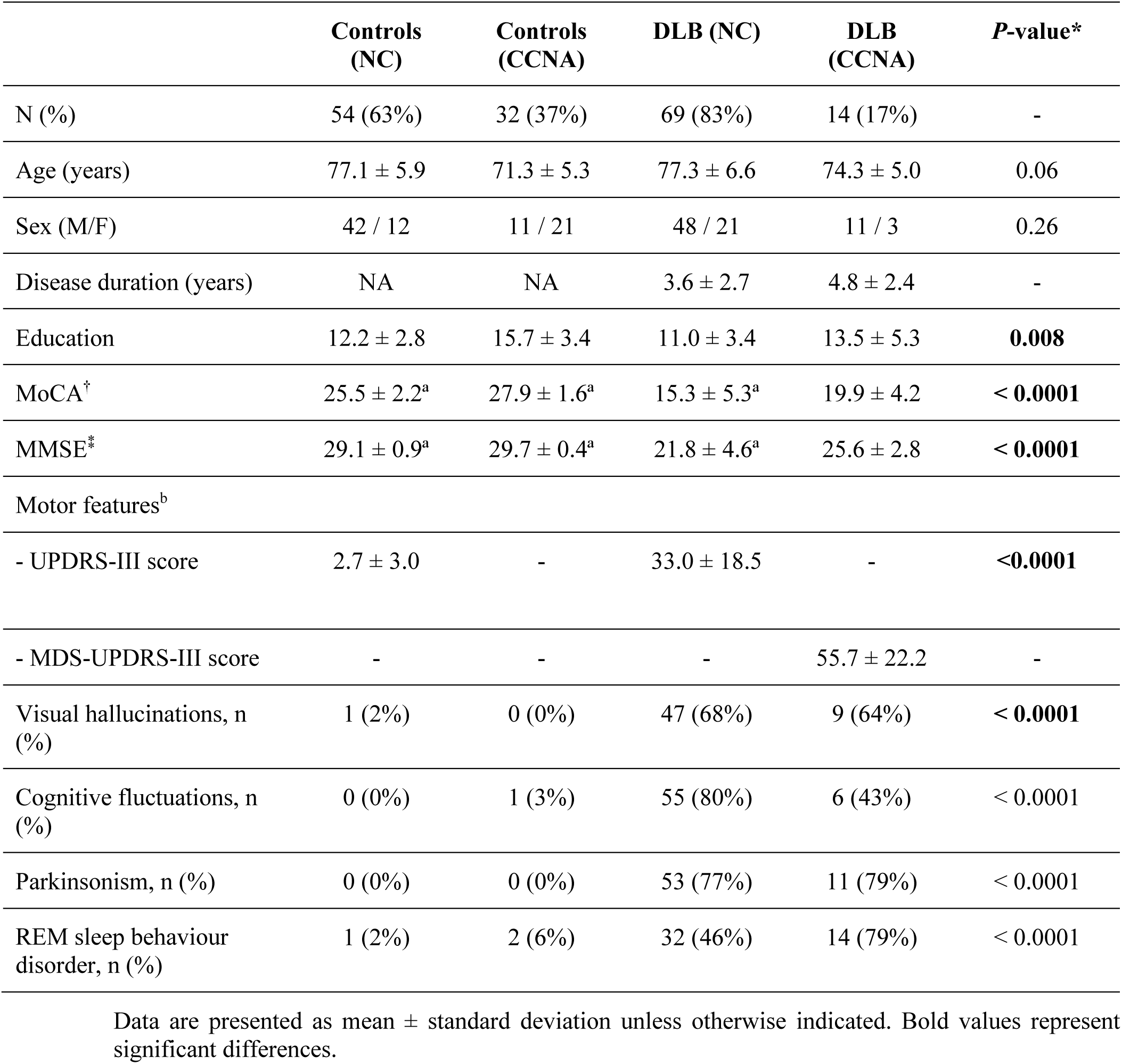
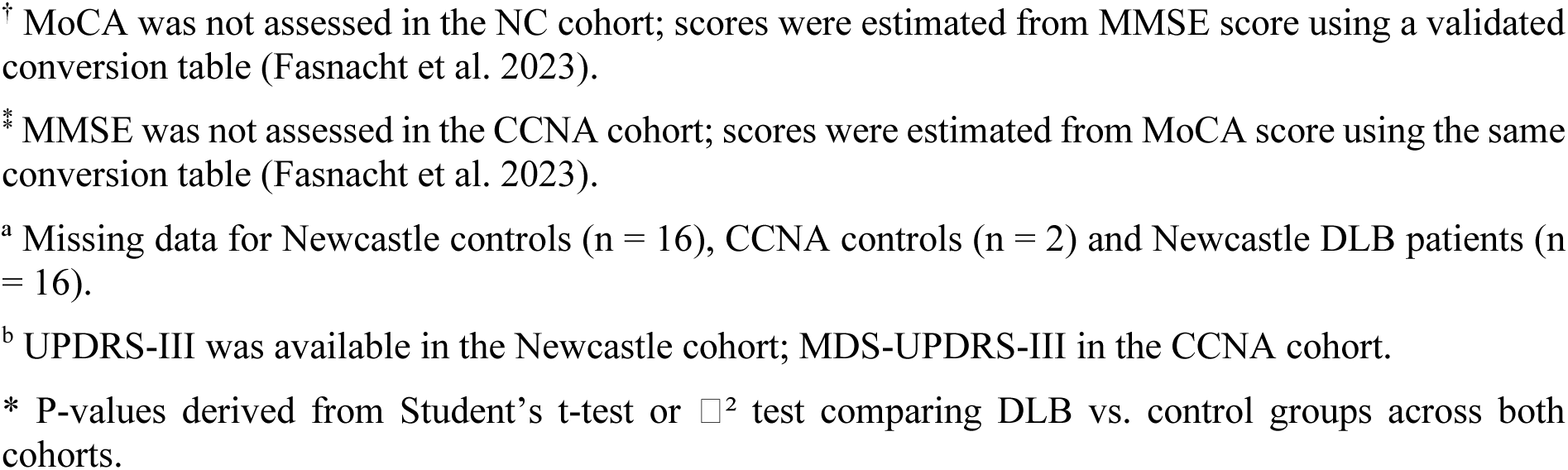
Demographic and clinical characteristics of DLB patients and controls.

|  | **Controls**<br>**(NC)** | **Controls**<br>**(CCNA)** | **DLB (NC)** | **DLB**<br>**(CCNA)** | **P-value\*** |
| --- | --- | --- | --- | --- | --- |
| N (%) | 54 (63%) | 32 (37%) | 69 (83%) | 14 (17%) | - |
| Age (years) | $77.1 \pm 5.9$ | $71.3 \pm 5.3$ | $77.3 \pm 6.6$ | $74.3 \pm 5.0$ | 0.06 |
| Sex (M/F) | 42 / 12 | 11 / 21 | 48 / 21 | 11 / 3 | 0.26 |
| Disease duration (years) | NA | NA | $3.6 \pm 2.7$ | $4.8 \pm 2.4$ | - |
| Education | $12.2 \pm 2.8$ | $15.7 \pm 3.4$ | $11.0 \pm 3.4$ | $13.5 \pm 5.3$ | **0.008** |
| MoCA<sup>†</sup> | $25.5 \pm 2.2^a$ | $27.9 \pm 1.6^a$ | $15.3 \pm 5.3^a$ | $19.9 \pm 4.2$ | **< 0.0001** |
| MMSE<sup>*</sup> | $29.1 \pm 0.9^a$ | $29.7 \pm 0.4^a$ | $21.8 \pm 4.6^a$ | $25.6 \pm 2.8$ | **< 0.0001** |
| Motor features<sup>b</sup> |  |  |  |  |  |
| - UPDRS-III score | $2.7 \pm 3.0$ | - | $33.0 \pm 18.5$ | - | **<0.0001** |
| - MDS-UPDRS-III score | - | - | - | $55.7 \pm 22.2$ | - |
| Visual hallucinations, n (%) | 1 (2%) | 0 (0%) | 47 (68%) | 9 (64%) | **< 0.0001** |
| Cognitive fluctuations, n (%) | 0 (0%) | 1 (3%) | 55 (80%) | 6 (43%) | < 0.0001 |
| Parkinsonism, n (%) | 0 (0%) | 0 (0%) | 53 (77%) | 11 (79%) | < 0.0001 |
| REM sleep behaviour disorder, n (%) | 1 (2%) | 2 (6%) | 32 (46%) | 14 (79%) | < 0.0001 |

Data are presented as mean ± standard deviation unless otherwise indicated. Bold values represent significant differences.
† MoCA was not assessed in the NC cohort; scores were estimated from MMSE score using a validated conversion table (Fasnacht et al. 2023).
\* MMSE was not assessed in the CCNA cohort; scores were estimated from MoCA score using the same conversion table (Fasnacht et al. 2023).
<sup>a</sup> Missing data for Newcastle controls (n = 16), CCNA controls (n = 2) and Newcastle DLB patients (n = 16).
<sup>b</sup> UPDRS-III was available in the Newcastle cohort; MDS-UPDRS-III in the CCNA cohort.
\* P-values derived from Student's t-test or $\chi^2$ test comparing DLB vs. control groups across both cohorts.

### Posterior-predominant cortical changes in DLB

We first characterized cortical thickness, surface area, and volume in patients with DLB relative to healthy controls using surface-based morphometry. First, vertex-based analyses showed widespread reductions in cortical thickness, cortical volume, and to a lesser extent, surface area (Fig.2A). FDR-corrected region-wise analyses further complemented these results, revealing that 82.4% of cortical regions showed significant thinning, 85.3% exhibited volume loss, and 22.1% showed reductions in surface area (Fig.2B and Supplementary Table 5-7). Cortical change was most pronounced in posterior regions, with reductions in cortical thickness and volume affecting the parietal, temporal, and occipital cortex, and, to a lesser degree the frontal cortex (Fig.2B, Supplementary Table 5 and 6). In contrast, reductions in surface area were more restricted and predominantly found in the parietal cortex (Fig.2B, Supplementary Table 7). No significant associations were found between regional cortical thinning and MoCA (range of regional β-coefficient estimates between -0.100 and 1.53, *P*_FDR_ > 0.05) and MDS-UPDRS-III scores (range of regional β-coefficient estimates between -6.75 and 3.59, *P*_FDR_ > 0.05) within the DLB group. Together, these findings align with previous studies showing that DLB is characterized by widespread cortical degeneration with a distinct posterior predominance.^9, 55, 56^

**Figure 2.**
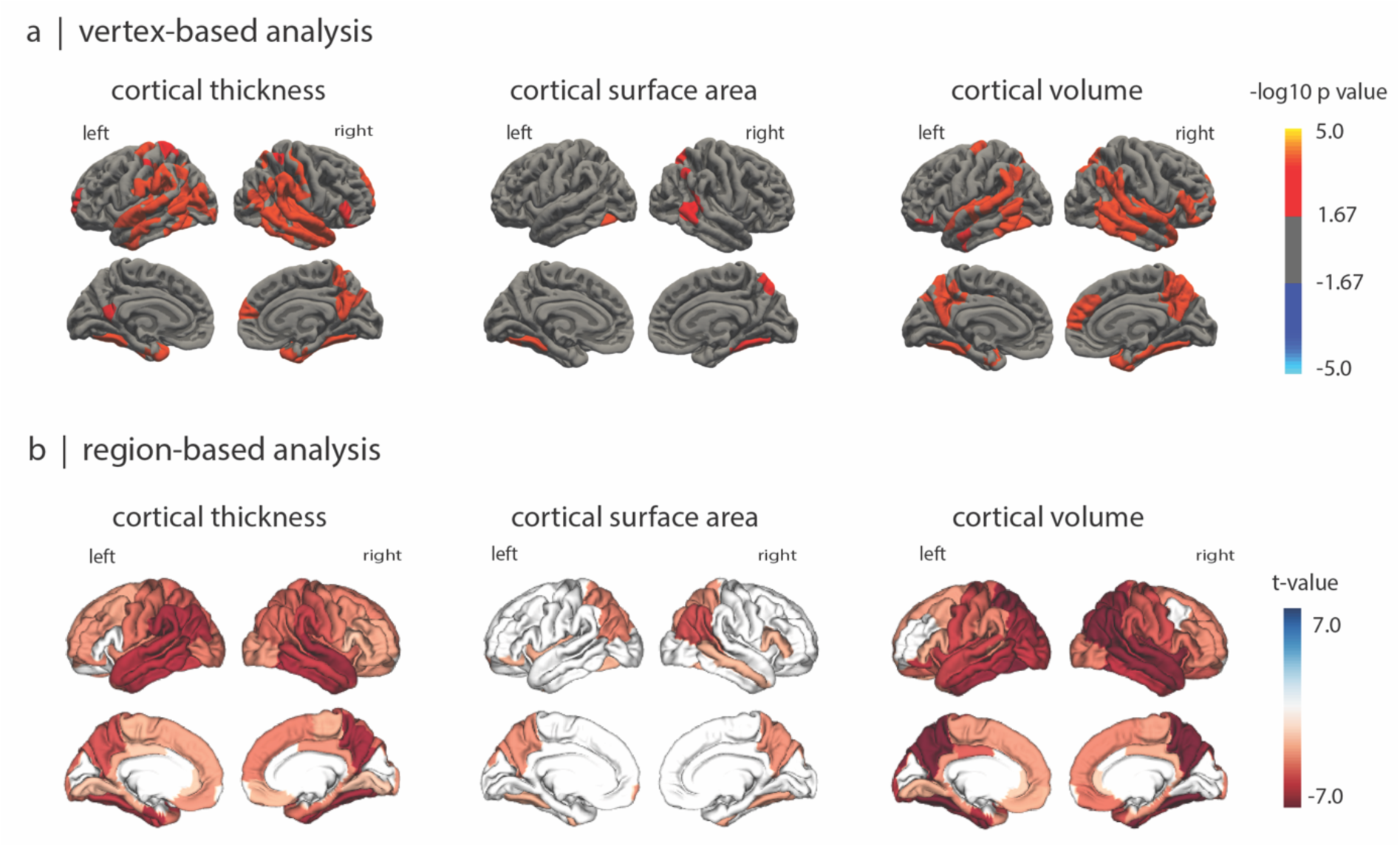
Patterns of cortical changes in DLB. **(a)** Vertex-wise surface analyses of cortical thickness, surface area, and cortical volume comparing DLB patients and controls. Colored surface maps show the -log10 P-values from vertex-wise GLMs implemented in FreeSurfer, controlling for age and site (and for eTIV in surface area and volume analyses). Red areas reflect lower values in DLB patients compared to controls. **(b)** t-value maps showing region-based ANOVA comparisons on ComBat-corrected W-scores, i.e., adjusted for age, sex, and site (and eTIV in surface area and volume models). Negative t-values (blue) reflect reduced morphometric values in the DLB group. Regions without significant group differences after FDR are shown in white. ANOVA, analysis of variance; DLB, dementia with Lewy bodies; eTIV, estimated total intracranial volume; FDR = false discovery rate.

### Cortical changes in DLB align with normative gene expression patterns

Prior evidence has shown that gene expression profiles may contribute to regional vulnerability in PD, iRBD, and multiple system atrophy, particularly through mitochondrial pathways.^22, 57-59^ Here, we investigated whether cortical changes in DLB follow a similar pattern of molecularly shaped vulnerability by testing its spatial correspondence with normative gene expression profiles in the healthy brain and identifying enriched biological processes among the contributing genes.

Partial least squares regression identified two significant LVs capturing shared covariance between regional cortical thinning in DLB and gene expression. LV1 and LV3 explained 35.6% and 22.3% of the covariance in cortical thickness (spatial null models: LV1: 10.9%, P_spatial_ = 0.0014; LV3: 7.9%, *P*_spatial_ = 0.0006) and random null models (LV1: 8.5%, *P*_random_ < 0.0001; LV3: 8.5%, *P*_random_ = 0.0002) (Supplementary Fig.1A). Although LV2 accounted for 17.3% of covariance and reached significance against random permutations (*P*_random_ = 0.027), it did not survive correction against spatial null models (*P*_spatial_ = 0.05) (Supplementary Fig.1A), suggesting its association was driven by spatial autocorrelation. The regional weights from LV1 that were correlated with MRI-derived cortical thickness W-scores (r = 0.60, *P* < 0.001) indicated that regions showing greater thickness reduction in DLB also had higher expression of negatively weighted genes (Supplementary Fig.1B). This spatial coupling was strongest in lateral temporo-parietal regions (Supplementary Fig.1C). Results for LV3 are shown in Supplementary Fig.2, similarly showing the strongest spatial coupling in the lateral temporo-parietal region. These findings suggest that the regional pattern of decreased cortical thickness in DLB reflects occurs in regions with distinct gene expression patterns in the healthy brain, highlighting a transcriptomic basis for selective vulnerability of regions in DLB.

### Genes associated with DLB cortical thinning are enriched for synaptic and mitochondrial processes

To identify biological functions among genes that were overexpressed in regions with cortical thinning in DLB, we performed gene set enrichment analysis on the bootstrapped gene weights from the imaging transcriptomics analysis and tested their enrichment across curated gene sets related to biological processes and cellular components.

This analysis revealed significant enrichment for 23 biological processes, with the strongest associations involving mitochondrial function and synaptic transmission (Fig.3A, Table 2). Among mitochondrial-related terms, the most enriched were electron transport chain (normalized enrichment score [NES] = -2.65, *P*_FDR_ < 0.0001), NADH dehydrogenase complex assembly (NES = -2.55, *P*_FDR_ < 0.0001), protein localization to the mitochondrion (NES = - 2.33, *P*_FDR_ = 0.0002), energy derivation by oxidation of organic compounds (NES = -2.21, *P*_FDR_ = 0.0006), and mitochondrion organization (NES = -2.18, *P*_FDR_ = 0.0006). Synaptic processes were also significantly enriched, including vesicle-mediated transport (NES: -2.59, *P*_FDR_ < 0.0001), regulation of trans-synaptic signalling (NES = -2.32, *P*_FDR_ = 0.0002), postsynaptic chemical transmission (NES = -2.20, *P*_FDR_ = 0.0006), and regulation of postsynaptic neurotransmitter receptor levels (NES = -2.18, *P*_FDR_ = 0.0006). Notably, of these terms, the *SNCA* gene (i.e., encoding alpha-synuclein) was implicated in 3 of the 4 enriched synaptic terms and 3 of the 5 mitochondrial terms. At the cellular component level, negatively weighted genes were enriched in mitochondrial structures such as the respirasome (NES = - 2.71, *P*_FDR_ < 0.0001), oxidoreductase complex (NES = -2.56, *P*_FDR_ < 0.0001), mitochondrial protein-containing complex (NES = -2.55, *P*_FDR_ < 0.0001), mitochondrial inner membrane (NES = -2.34, *P*_FDR_ < 0.0001), and cytochrome complex (NES = -2.19, *P*_FDR_ = 0.0003). Synaptic structures were also significantly represented, including the neuron-to-neuron synapse (NES = -2.32, *P*_FDR_ < 0.0001), glutamatergic synapse (NES = -2.29, *P*_FDR_ < 0.0001), postsynaptic specialization (NES = -2.28, P_FDR_ < 0.0001), and presynaptic active zone (NES = -2.02, *P*_FDR_ = 0.002) (Table 2). To test convergence with Lewy body dementia genetics, we assessed whether GWAS-prioritized Lewy body dementia genes were overexpressed among the negatively weighted genes linked to DLB cortical thinning. Of the 69 Lewy body dementia risk genes reported in previously referenced gene-level GWAS study,^49^ 57 had expression values in our transcriptomic dataset and 13 were overexpressed. The most strongly associated genes were *FAM171A2* (key regulator of progranulin expression) and *SNCA* (bootstrap ratio < -4.0, approximate two-tailed P values of *P* < 0.000063) followed by *NSF*, *CLPTM1*, *MAPT*, *GAK* (bootstrap ratio < -3.0, *P* < 0.0027), and *STX4*, *PPP1R37*, *CLU*, *APOC1*, *FGFRL1*, *UBQLN4*, and *EFNA3* (bootstrap ratio < -2.0, *P* < 0.046) (Fig.3B).

**Figure 3.**
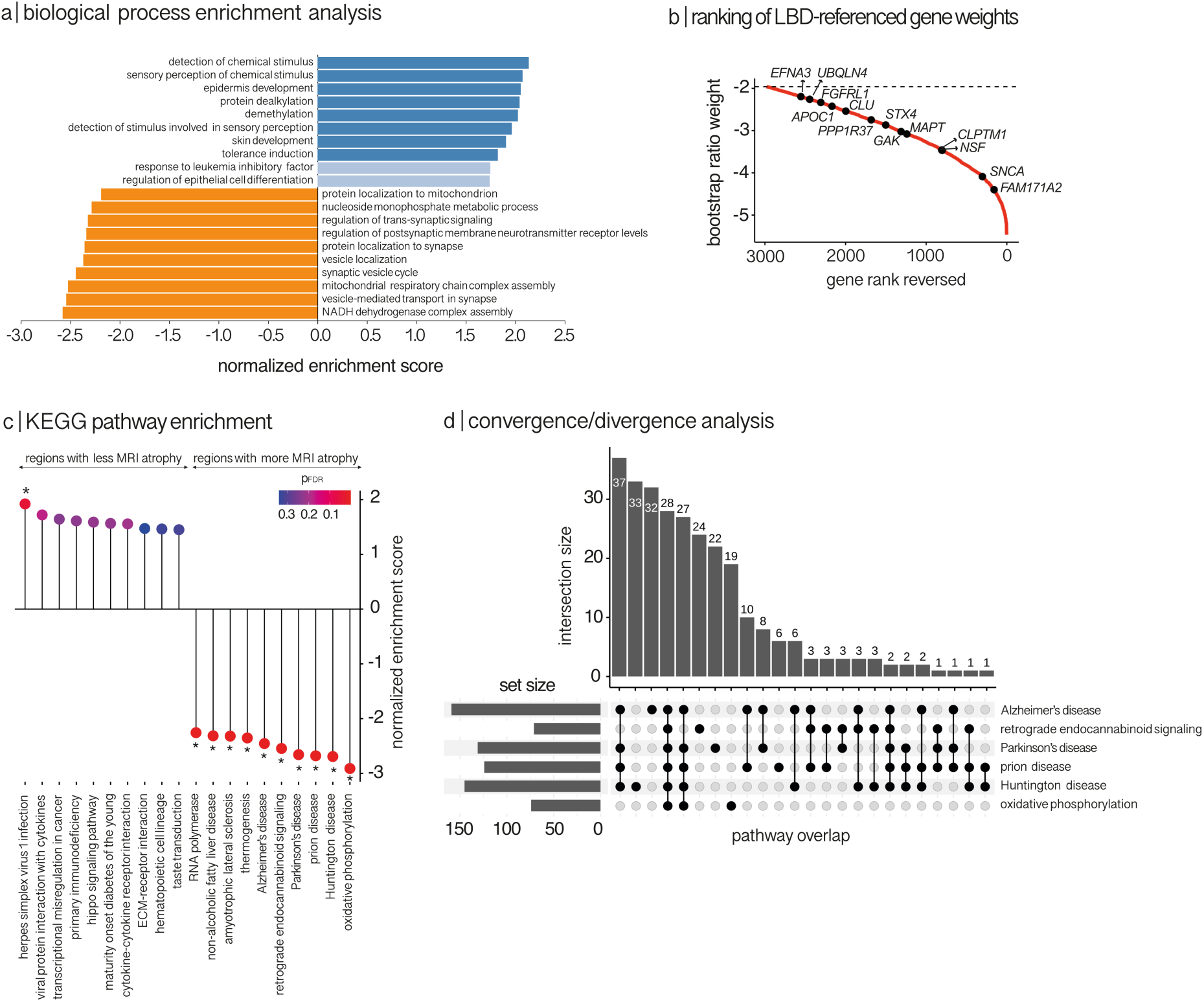
Functional enrichment of genes associated with cortical thickness in DLB. **(a)** GO biological process enrichment analysis of LV1. Bar plot showing the top 10 GO terms enriched among negatively weighted genes (more highly expressed in regions with greater cortical thinning; orange) and positively weighted genes (regions with relatively preserved thickness; blue). Bars indicate normalized enrichment score; lighter shading denotes terms not surviving FDR correction. **(b)** LBD-referenced genes among negatively weighted genes (regions showing change in DLB). Genes previously implicated in LBD by a gene-level GWAS were mapped to our transcriptomic set and those with bootstrap ratio below -1.96 (outside the central 95% of a standard normal) are plotted. **(c)** KEGG pathway enrichment. Dot plot showing KEGG pathways enriched in negatively and positively weighted genes. The x-axis lists pathways; the y-axis shows normalized enrichment scores. Colour intensity reflects statistical significance (P_FDR_-value). Asterisks indicate pathways surviving FDR correction. **(d)** UpSet plot illustrating overlap among KEGG pathways enriched in negatively weighted genes (greater cortical thinning). Vertical bars represent the number of shared genes between pathway sets; horizontal bars indicate the size of each individual pathway. Pathways related to neurodegenerative diseases exhibited substantial convergence. DLB, dementia with Lewy bodies; GO, Gene Ontology, FDR, false discovery rate; KEGG, Kyoto Encyclopaedia of Genes and Genomes; LBD, Lewy body dementia; LV, latent variable; NADH, nicotinamide adenine dinucleotide, reduced form; NES, normalized enrichment score.

**Table 2.** Biological processes and cellular components enriched in genes associated with cortical thickness in DLB.

| GO identifier | GO term | Gene set size | Leading edge genes | ES | NES | PFDR-value |
| --- | --- | --- | --- | --- | --- | --- |
| **Biological processes** |  |  |  |  |  |  |
| **Negatively weighted genes (overexpressed in regions with decreased thickness)** |  |  |  |  |  |  |
| GO:0022900 § | electron transport chain | 146 | 83 | -0.50 | -2.65 | < 0.0001 |
| GO:0099003 § | vesicle-mediated transport in synapse | 205 | 70 | -0.47 | -2.59 | < 0.0001 |
| GO:0010257 § | NADH dehydrogenase complex assembly | 54 | 28 | -0.59 | -2.55 | < 0.0001 |
| GO:0070585 <sup>§</sup> | protein localization to mitochondrion | 122 | 61 | -0.46 | -2.33 | 0.0002 |
| GO:0099177 <sup>§</sup> | regulation of trans-synaptic signalling | 425 | 169 | -0.38 | -2.32 | 0.0002 |
| GO:1902414 <sup>§</sup> | protein localization to cell junction | 103 | 45 | -0.45 | -2.21 | 0.007 |
| GO:0015980 <sup>§</sup> | energy derivation by oxidation of organic compounds | 280 | 140 | -0.38 | -2.21 | 0.0006 |
| GO:0099565 | chemical synaptic transmission, postsynaptic | 94 | 41 | -0.45 | -2.20 | 0.0006 |
| GO:0099072 <sup>§</sup> | regulation of postsynaptic membrane neurotransmitter receptor levels | 82 | 29 | -0.46 | -2.18 | 0.0006 |
| GO:0007005 <sup>§</sup> | mitochondrion organization | 439 | 188 | -0.36 | -2.18 | 0.0006 |
| GO:0070050 <sup>§</sup> | neuron cellular homeostasis | 55 | 27 | -0.51 | -2.21 | 0.0007 |
| GO:1902600 <sup>§</sup> | proton transmembrane transport | 114 | 59 | -0.40 | -2.04 | 0.004 |
| GO:0048499 | synaptic vesicle membrane organization | 27 | 16 | -0.55 | -2.04 | 0.003 |
| GO:0009141 <sup>§</sup> | nucleoside triphosphate metabolic process | 223 | 89 | -0.37 | -2.03 | 0.003 |
| GO:1901293 <sup>§</sup> | nucleoside phosphate biosynthetic process | 240 | 111 | -0.36 | -2.03 | 0.003 |
| GO:0072522 <sup>§</sup> | purine-containing compound biosynthetic process | 216 | 99 | -0.35 | -1.96 | 0.006 |
| GO:0050808 <sup>§</sup> | synapse organization | 440 | 143 | -0.32 | -1.95 | 0.006 |
| GO:0019693 § | ribose phosphate metabolic process | 396 | 172 | -0.33 | -1.94 | 0.006 |
| GO:0035249 | synaptic transmission, glutamatergic | 97 | 38 | -0.39 | -1.92 | 0.008 |
| GO:0050803 § | regulation of synapse structure or activity | 225 | 79 | -0.35 | -1.92 | 0.008 |
| GO:0043161 § | proteasome-mediated ubiquitin-dependent protein catabolic process | 412 | 141 | -0.29 | -1.76 | 0.022 |
| GO:0072594 § | establishment of protein localization to organelle | 400 | 153 | -0.29 | -1.75 | 0.022 |
| GO:0006887 § | exocytosis | 289 | 88 | -0.29 | -1.67 | 0.037 |
| ***Positively weighted genes (overexpressed in regions with increased thickness)*** |  |  |  |  |  |  |
| GO:0070828 § | heterochromatin organization | 88 | 42 | 0.52 | 2.46 | < 0.0001 |
| GO:1902275 | regulation of chromatin organization | 29 | 14 | 0.57 | 2.11 | 0.005 |
| GO:0007606 § | sensory perception of chemical stimulus | 107 | 54 | 0.43 | 2.09 | 0.006 |
| GO:0009593 § | detection of chemical stimulus | 97 | 51 | 0.43 | 2.04 | 0.008 |
| GO:0070269 § | pyroptosis | 25 | 15 | 0.58 | 2.01 | 0.009 |
| GO:0040029 | epigenetic regulation of gene expression | 132 | 50 | 0.39 | 1.97 | 0.014 |
| GO:0035329 | hippo signalling | 35 | 17 | 0.50 | 1.95 | 0.014 |
| ***Cellular components*** |  |  |  |  |  |  |
| ***Negatively weighted genes (overexpressed in regions with decreased thickness)*** |  |  |  |  |  |  |
| GO:0070469 § | respirasome | 86 | 59 | -0.57 | -2.71 | < 0.0001 |
| GO:1990204 § | oxidoreductase complex | 107 | 70 | -0.51 | -2.56 | < 0.0001 |
| GO:0098798 § | mitochondrial protein- | 273 | 167 | -0.45 | -2.55 | < 0.0001 |

|  | containing<br>complex |  |  |  |  |  |
| --- | --- | --- | --- | --- | --- | --- |
| GO:0005743 § | mitochondrial<br>inner membrane | 431 | 207 | -0.39 | -2.34 | < 0.0001 |
| GO:0098984 § | neuron to neuron<br>synapse | 339 | 157 | -0.39 | -2.32 | < 0.0001 |
| GO:0098978 § | glutamatergic<br>synapse | 390 | 174 | -0.38 | -2.29 | < 0.0001 |
| GO:0099572 § | postsynaptic<br>specialization | 320 | 143 | -0.39 | -2.28 | < 0.0001 |
| GO:0070069 § | cytochrome<br>complex | 33 | 22 | -0.56 | -2.19 | 0.0003 |
| GO:0048786 § | presynaptic<br>active zone | 71 | 36 | -0.44 | -2.02 | 0.002 |
| GO:0044309 § | neuron spine | 165 | 63 | -0.37 | -1.98 | 0.002 |

List of genes significantly enriched GO biological process and cellular component terms among genes associated with LV1 after FDR correction using WebGestalt. Gene terms are ranked by decreasing NES. For cellular components, there were no significant terms in positively weighted genes.
§ GO process terms also found with Panther.

In contrast, genes positively weighted on LV1 (i.e., those more highly expressed in regions with preserved or increased cortical thickness) were enriched for 7 biological processes, including stimulus detection, chromatin organization, and intracellular transport (Fig.3A, Table 2). No significant cellular components were associated with increased thickness (Table 2).

When using PANTHER for validating enrichment patterns, we found largely similar results for LV1 (Table 2). Enrichment analyses for LV3 revealed similar enrichment patterns as LV1, suggesting that both gene-thickness LVs pinpoint similar processes (Supplementary Table 8).

While a significant LV was found for surface area, no enrichment of synaptic or mitochondrial pathways was found in regions showing surface area reductions; instead, enrichment was only observed for increased surface area and included immune and metabolic pathways. Cortical volume, a composite of thickness and surface area, showed a gene expression pattern closely aligned with cortical thinning, with enrichment of synaptic signalling and mitochondrial processes across both significant latent variables (Supplementary Fig.3, Supplementary Table 9). These findings demonstrate that regions showing cortical thinning in DLB overexpress genes related to mitochondrial and synaptic functions, whereas surface area alterations may involve distinct biological processes.

### Neurodegenerative pathway enrichment in regions vulnerable to cortical thinning in DLB

We next sought to determine whether genes overexpressed in regions showing cortical thinning in DLB also participate in canonical neurodegenerative disease pathways. Cortical thickness was chosen as the metric of interest for subsequent analyses because reductions in cortical surface area did not show significant transcriptomic enrichment, whereas cortical volume mathematically represents the combination of cortical thickness and surface area.^60, 61^ Thickness and surface area are distinct geometric constituents of cortical volume, with partly different genetic and developmental determinants.^62, 63^ Cortical thickness is also the most widely studied surface-based metric in neurodegeneration and unlike surface area, does not scale directly with brain size,^64^ thereby reducing unwanted interindividual variance when assessing neurodegenerative effects on cortical structure. Here, our analysis aimed to contextualize the molecular mechanisms underlying DLB-related cortical thinning within well-characterized pathways implicated in related disorders, particularly PD and AD, which overlap with DLB pathophysiology. Using the KEGG knowledge base with 6,290 gene IDs annotated, we found that the most enriched pathway was oxidative phosphorylation (hsa00190; 74 of 112 [66.1%] leading-edge genes), followed by neurodegenerative and signalling pathways, including Huntington’s disease (hsa0516; 145/226 [64.2%] genes), prion disease (hsa05020; 124/233 [53.2%] genes), Parkinson’s disease (hsa05012; 131/236 [55.5%] genes), retrograde endocannabinoid signalling (hsa04723; 71/129 [55.0%] genes), and Alzheimer’s disease (hsa05010; 159/332 [47.9%] genes) (Fig.3C). Given the known overlap between these pathways, we assessed the convergence and divergence of contributing genes. A set of 28 genes was shared across all six pathways (Fig.3D). These genes were predominantly related to mitochondrial function, particularly subunits of the NADH:ubiquinone oxidoreductase (NDUF) family (Fig.4A). Over-representation analysis of this shared gene set confirmed enrichment for mitochondrial complex I assembly and electron transport chain function (Fig.4B and Supplementary Table 10).

**Figure 4.**
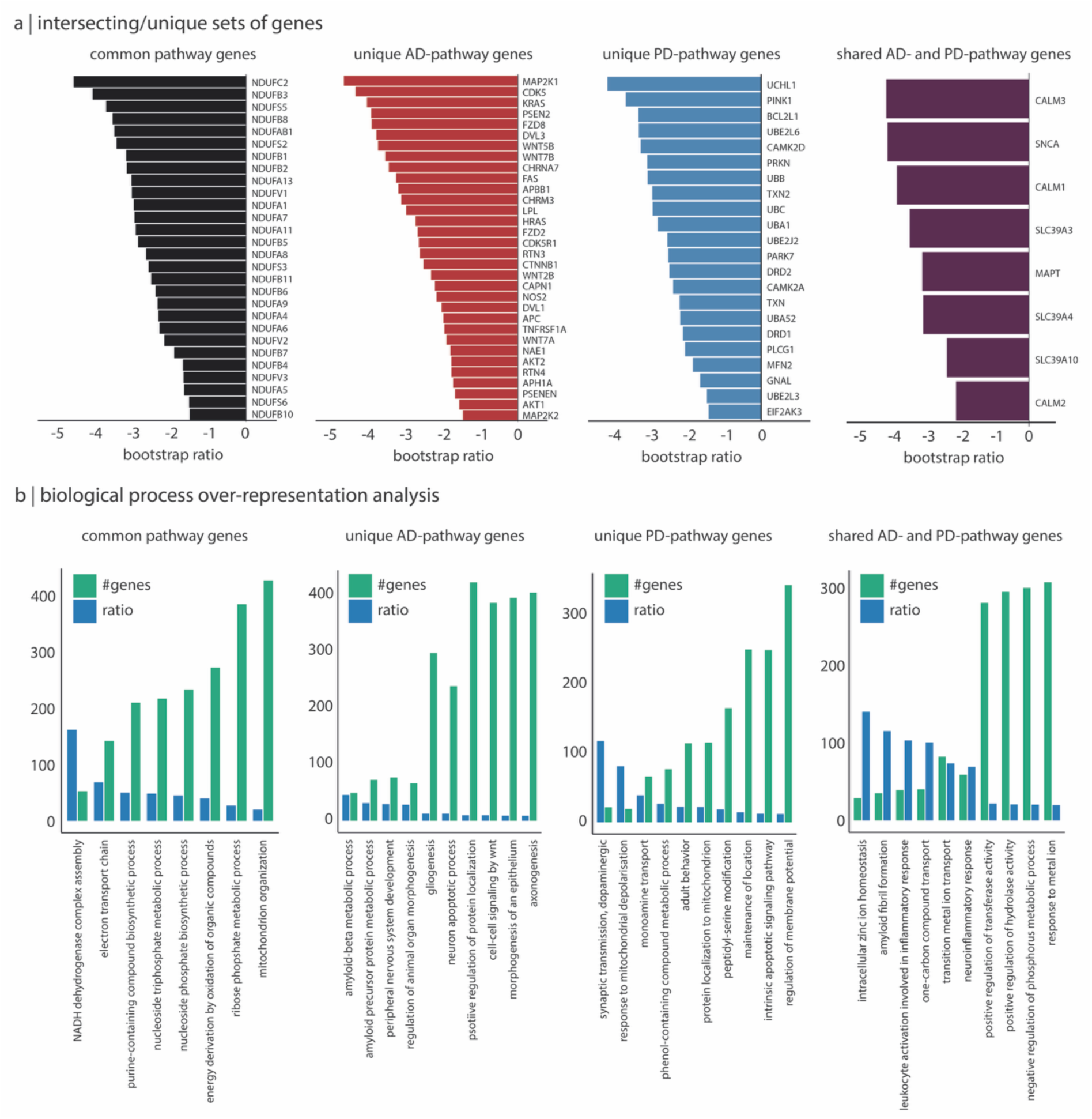
Shared and unique genes and associated biological processes across pathways enriched in regions showing cortical thinning in DLB. **(a)** Genes identified from pathway enrichment analysis of genes overexpressed in regions with DLB-related cortical thinning. Shown are genes common to all six enriched KEGG pathways, genes unique to the AD or PD pathways, and genes shared between the AD and PD pathways. Genes are ordered by decreasing bootstrap ratio. Over-representation analysis of GO biological processes for each gene group. Green bars represent the number of genes associated with each term; blue bars indicate the enrichment ratio. AD, Alzheimer’s disease; DLB, dementia with Lewy bodies; GO, Gene Ontology; KEGG, Kyoto Encyclopaedia of Genes and Genomes; PD, Parkinson’s disease.

We also identified pathway-specific gene sets. A set of 32 genes overexpressed in regions with DLB-related cortical thinning were unique to the AD pathway (Fig.3D). The top contributors included *MAP2K1*, *CDK5*, *KRAS*, and *PSEN2*, all showing bootstrap ratio weights below -3.8 (*P* < 0.0001) (Fig.4A). These genes were specifically enriched for amyloid-beta and amyloid precursor protein metabolism (Fig.4B and Supplementary Table 10). Similarly, 22 genes were uniquely annotated within the PD pathway (Fig.3D), including *UCHL1*, *PINK1*, *BCL2L1*, *UBE2L6*, *CAMK2D*, and *PRKN*, with bootstrap weights below -2.9 (*P* < 0.005) (Fig.4A). These genes were enriched for dopaminergic synaptic transmission and response to mitochondrial depolarization (Fig.4B and Supplementary Table 10). A smaller subset of 8 genes was shared exclusively between the AD and PD pathways (Fig.3D), included canonical neurodegeneration-related genes *SNCA* and *MAPT* (Fig.4A), and was enriched for processes such as intracellular zinc ion homeostasis and amyloid fibril formation (Fig.4B and Supplementary Table 10).

We also identified a group of 90 genes overexpressed in regions with cortical thinning in DLB that were not included in the KEGG AD or PD pathways. These genes may underly mechanisms of regional cortical vulnerability more specific to DLB. Over-representation analysis revealed strong enrichment for the GABAergic signalling pathway (enrichment ratio = 26.6, *P*_FDR_ < 0.0005), including multiple GABA_A_ receptor subunits (α-3, β-1, β-3, ε, and γ-1), as well as for neuron cellular homeostasis (enrichment ratio = 26.6, *P*_FDR_ < 0.0001). Additional enriched biological processes included proton transmembrane transport, pH regulation, vesicle-mediated synaptic transport, monoatomic anion transport, and receptor-mediated endocytosis (Fig.5A and Supplementary Table 11). Together, these findings highlight that while DLB-related cortical thinning overlaps with AD and PD pathways, it also implicates distinct transcriptomic vulnerabilities, particularly related to GABAergic transmission.

**Figure 5.**
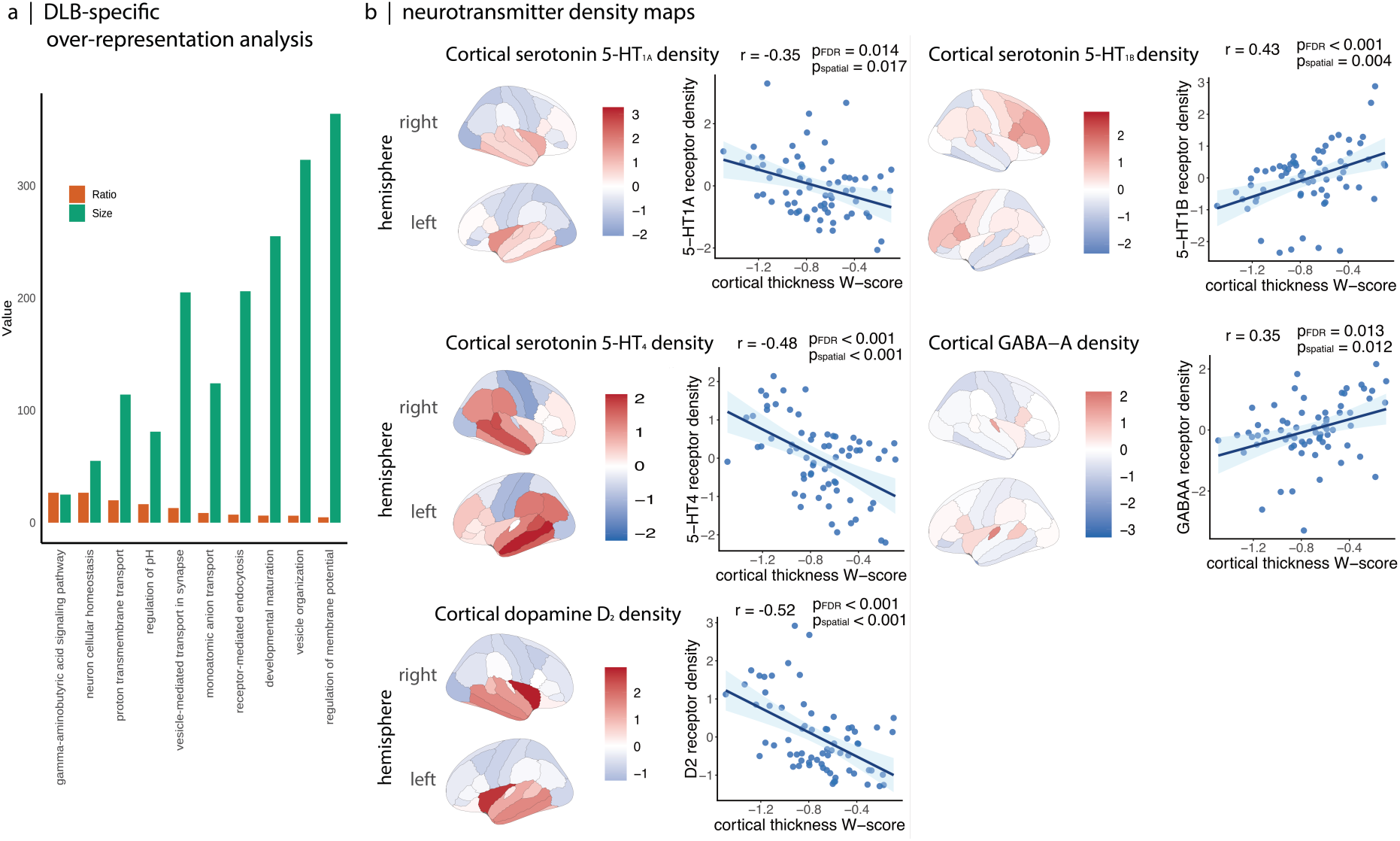
DLB-specific over-representation analysis and neurotransmitter density maps in relation to cortical thinning in DLB. **(a)** Over-representation results of biological processes enriched in the genes not part of KEGG AD and PD pathways. Each bar represents a significantly enriched biological process (PFDR < 0.05). The green bar corresponds to the number of genes within the gene set, and the orange bar indicates the bootstrap ratio weight, reflecting the robustness of the enrichment. **(b)** Brain surface renderings showing the z-scored spatial distribution of neurotransmitter receptor densities significantly correlated with cortical thickness W-scores in DLB. Scatterplots display Spearman correlations between z-scored receptor density and cortical thickness W-scores, along with P-values derived from spatial permutation testing. AD, Alzheimer’s disease; FDR, false discovery rate; KEGG, Kyoto Encyclopaedia of Genes and Genomes; PD, Parkinson’s disease; DLB, dementia with Lewy bodies.

### Brain specificity of DLB cortical thinning genes

To further characterize the neurodegenerative relevance of the DLB-specific genes, we investigated whether they were preferentially expressed in brain tissue compared to peripheral organs. Using expression data from GTEx across 68 tissue types (Supplementary Table 2 and Fig.6A), including 13 brain regions, we found that 42 out of the 90 genes (47%) enriched in regions vulnerable to DLB cortical thinning, but not associated with AD and PD pathways, were significantly overexpressed in the brain (Fig.6B and Supplementary Table 12). Functional enrichment analysis revealed that this brain-expressed gene set was significantly associated with regulation of trans-synaptic signalling (enrichment ratio = 2.05, *P*_FDR_ = 0.0089) (Fig.6C and Supplementary Table 13), suggesting a potential role of these brain-expressed genes in inter-regional neuronal communication and potential prion-like spread.

**Figure 6.**
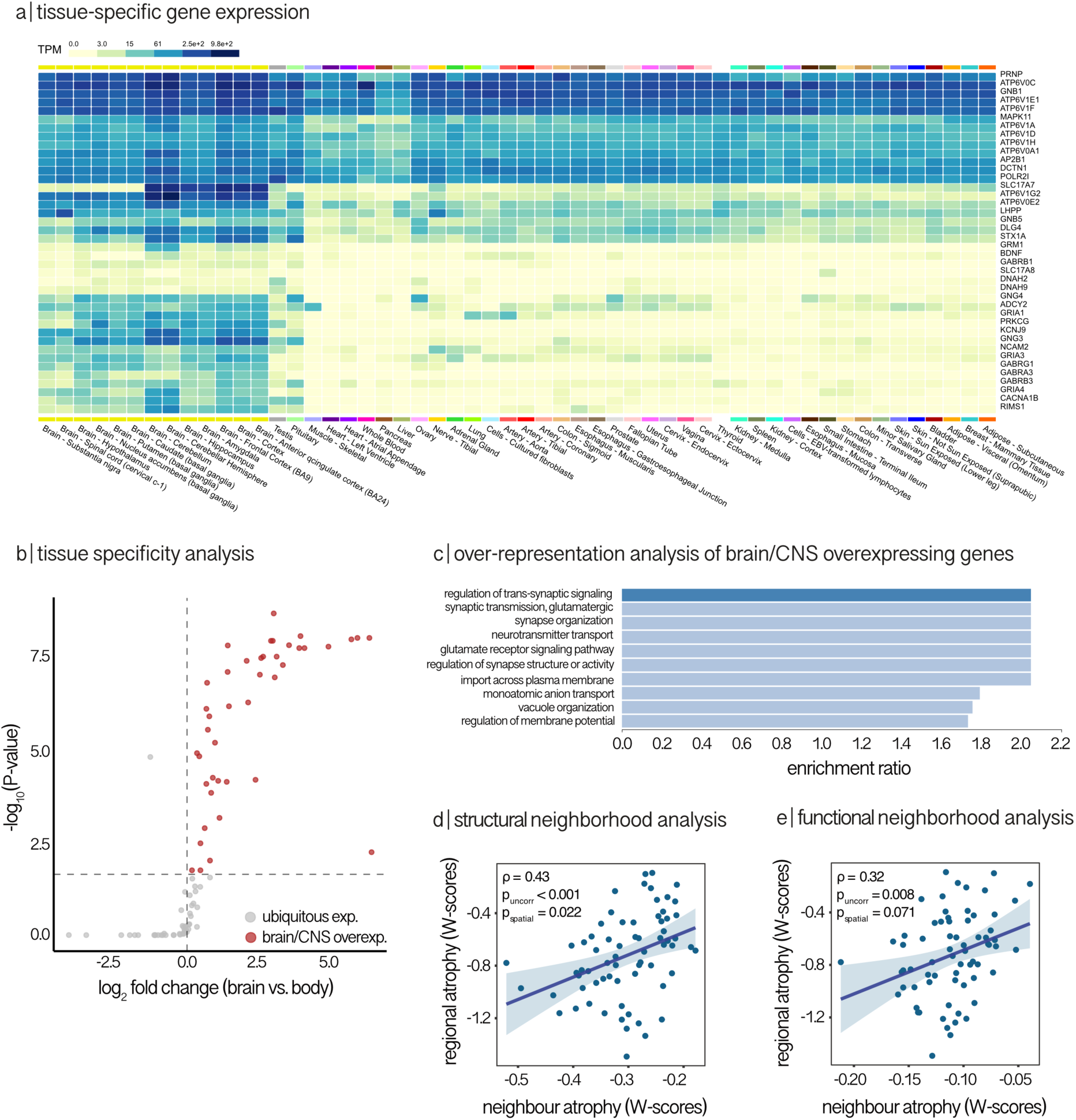
Brain-enriched expression of DLB-associated genes and network constraints on cortical thinning. **(a)** Matrix showing TPM expression values for the 42 DLB-associated genes identified as significantly enriched in brain tissue across 68 tissue types from the GTEx dataset. Brain tissues are shown on the left, followed by the non-brain tissues; darker blue indicates higher expression levels. **(b)** Volcano plot of tissue enrichment analysis for all 90 DLB-associated genes, showing tissue types with significantly increased expression (P_FDR_ < 0.05); genes enriched in brain tissues are highlighted in red. **(c)** Bar plots showing the top significantly enriched GO biological processes for the 42 brain-enriched genes, ranked by enrichment ratio; lighter bars indicate terms not surviving FDR correction. **(d-e)** Scatterplots showing the relationship between regional cortical atrophy in DLB and average atrophy in structurally and functionally connected neighbours, weighted by the respective connectivity matrices. DLB, dementia with Lewy bodies; FDR, false discovery rate; GO, Gene Ontology; GTEx, Genotype-Tissue Expression; TPM, transcript per million.

### Structural connectivity influences the cortical thinning pattern in DLB

Previous evidence indicates that misfolded pathological proteins may spread through the brain via prion-like mechanisms,^13, 65^ including in PD and iRBD, where MRI-derived neurodegeneration patterns correlate with in silico simulations of pathological protein propagation.^20, 21, 59, 66^ Consistent with this framework, normative structural connectivity has been shown to constrain the propagation of pathology across brain networks.^20, 59^ The enrichment of trans-synaptic signalling processes among DLB-specific genes in the previous analysis further supports the relevance of this mechanism in DLB.

Here, we further investigated whether the spatial pattern of cortical thinning in DLB was shaped by the brain’s structural and functional connectome. A neighbourhood analysis revealed that regional cortical thinning in DLB significantly correlated with the degree of cortical thinning in each region’s structurally connected neighbours (ρ = 0.43, *P*_original_ < 0.001, *P*_spatial_ = 0.022) (Fig.6D). To determine whether functional co-activation patterns also contributed, we assessed the association between cortical thinning and the resting-state functional connectome. While a moderate correlation was observed (ρ = 0.32, *P*_original_ = 0.008), this association did not reach significance using spatial permutation testing (*P*_spatial_ = 0.071) (Fig.6E). These results indicate that cortical thinning in DLB is more strongly shaped by structural connectivity than by functional coupling, supporting a model in which disease-related processes may propagate along structural pathways between brain regions.

### Spatial correlation between cortical thinning and neurotransmitter systems

Synaptic pathology is a central neuropathological feature of major neurocognitive disorders, particularly DLB, where it contributes to synaptic dysfunction, synapse loss, and altered neurotransmission.^67-69^ Our gene enrichment analyses have implicated GABAergic processes among DLB-specific genes, consistent with evidence linking visual hallucinations to abnormal GABAergic transmission in posterior brain regions.^70^ However, whether cortical thinning in DLB converges spatially with the normative density of neurotransmitter receptors and transporters remains unclear. We therefore investigated spatial correlations between cortical thinning in DLB and PET- and SPECT-derived maps of neurotransmitter receptor and transporter density, including GABAergic markers (Fig.5B and Supplementary Table 14). Although GABA_A_ receptors were widely expressed across the cortex, their density was significantly lower in the regions most affected by cortical thinning in DLB (r = 0.36, *P*_FDR_ = 0.013, *P*_random_ = 0.0014, *P*_spatial_ = 0.012). In addition to GABAergic systems, we observed significant spatial associations with serotonergic and dopaminergic markers. Cortical thinning was more pronounced in regions with higher density of 5-HT_1A_ (r = -0.35, *P*_FDR_ = 0.014, *P*_random_ < 0.001, *P*_spatial_ = 0.017), 5-HT_4_ (r = -0.48, *P*_FDR_ < 0.001, *P*_random_ < 0.001, *P*_spatial_ < 0.001), and D2 (r = -0.52, *P*_FDR_ < 0.001, *P*_random_ < 0.001, *P*_spatial_ < 0.001) receptors. Conversely, regions with greater cortical thinning exhibited lower 5-HT_1B_ receptor density (r = 0.43, P_FDR_ = 0.002, *P*_random_ < 0.001, *P*_spatial_ = 0.004). No significant spatial correlations were found for acetylcholine, dopamine D1, noradrenaline, serotonin 5-HT_2A_ and 5-HT_6_ receptors, serotonin transporter 5-HTT, glutamate, endocannabinoid, opioid, histamine, or SV2A systems (*P*_FDR_ > 0.05). Combined high GABAergic signalling gene expression and low GABA_A_ (benzodiazepine-site availability) receptor density in regions vulnerable to DLB-related cortical thinning may reflect vulnerability of inhibitory circuitry in DLB.

## Discussion

In this study, we identified a distinct pattern of posterior-predominant cortical thinning in DLB, with most thinning occurring in the parietal, temporal, occipital, and posterior frontal cortices. Using imaging transcriptomics, we showed that this cortical thinning pattern overlaps with gene expression in the healthy brain, particularly genes involved in synaptic transmission and mitochondrial function. While many of these genes were shared with canonical neurodegenerative pathways implicated in PD and AD, we also identified a distinct subset of 90 genes related with neither pathway. These DLB-enriched genes were strongly associated with GABAergic signalling and cellular homeostasis, and nearly half were preferentially expressed in brain tissue, supporting their specificity to CNS function. Spatial correlations with neurotransmitter receptor density also implicated GABA_A_ receptor distribution in regions with cortical thinning. In addition, the cortical thinning pattern was constrained by structural connectivity, consistent with trans-neuronal propagation along structural pathways. Together, these findings suggest that DLB-related cortical thinning occurs in regions characterized by distinctive gene expression profiles, specific neurotransmitter receptor distributions, and structural connectivity patterns that may facilitate disease spread.

Our results align with a large body of literature showing that structural MRI reveals significant cortical changes in DLB, particularly in posterior brain regions.^6, 35, 56, 71-73^ These posterior changes are also seen in prodromal stages of DLB^74, 75^ and across imaging modalities, including occipital hypoperfusion on perfusion SPECT^76^ and the cingulate island sign, used to differentiate DLB from AD, on FDG-PET.^77^ Extending these observations, our imaging transcriptomics analyses revealed that the atrophied cortical regions in DLB are enriched for genes involved in mitochondrial structure and synaptic transmission. This supports established evidence implicating mitochondrial dysfunction in synucleinopathies. For instance, reduced complex I activity and impaired mitochondrial oxygen uptake have been reported in cortical samples from both DLB and PD patients, along with increased markers of oxidative stress.^78^ In the nucleus basalis of Meynert, a region severely affected in both manifest and prodromal DLB,^79-82^ reduced complex I expression (relative to mitochondrial mass) correlates with neuronal loss independently of Lewy body or amyloid pathology.^83^

These findings also converge with previous imaging transcriptomics studies in related neurodegenerative conditions. In PD, regions showing cortical thinning and longitudinal tissue deformation are those with high expression of mitochondrial and synaptic genes.^21^ Similarly in iRBD, a prodromal synucleinopathy with high conversion risk to DLB or PD,^19^ atrophy predicted DLB development^29, 55^ and preferentially emerges in structurally connected regions enriched for complex I-related gene expression.^22^ By contrast, AD exhibits a distinct pattern of atrophy in regions enriched for genes involved in protein folding and lipid metabolism, with *APOE* emerging as a key driver.^22^ Collectively, these disease-specific transcriptomic profiles reinforce the value of imaging transcriptomics in uncovering the molecular substrates of neurodegeneration as seen on MRI. In this study, overlaying Lewy body disease-related GWAS-prioritized genes onto our imaging transcriptomic map showed that the strongest associations in regions showing DLB-related cortical thinning were for *SNCA*, encoding alpha-synuclein, and *FAM171A2*, a mediator for alpha-synuclein fibril internalization.^84^

We further found that the genes associated with cortical thinning in DLB significantly overlapped with KEGG-defined pathways for both PD and AD using a data-driven analysis without prior assumptions. This supports the well-established notion that DLB lies biologically at the intersection of these two disorders.^28^ Previous studies have shown that the motor-dementia interval, which has been previously used to clinically differentiate DLB from PD dementia,^1^ is influenced by the burden of AD co-pathology. Individuals with higher levels of AD pathology tend to exhibit earlier cognitive symptoms, whereas those with minimal AD pathology more often show a motor-first progression.^85-87^ The biological heterogeneity is increasingly recognized in prodromal populations such as patients with iRBD, where plasma markers of AD pathology can predict conversion first to DLB rather than PD.^88^

Importantly, our study also identified a distinct set of 90 genes that were specifically enriched in regions vulnerable to cortical thinning in DLB but did not map onto existing KEGG pathways for either AD or PD. These DLB-enriched genes were primarily involved in GABAergic signalling and cellular homeostasis. Notably, regions with the greatest cortical thinning demonstrated high expression of GABAergic signalling genes but low GABA_A_ receptor density in the healthy brain. Importantly, PET and genes probe related but not identical facets of inhibitory circuitry: PET result reflects GABA_A_ receptor availability while transcriptomic results reflect the broader program spanning synthesis, packaging, transport, receptor subunits, and scaffolding related to GABAergic signalling in the brain. This interplay between transcription and availability may contribute to the selective vulnerability associated with brain neurodegeneration in DLB. Supporting this, reduced occipital GABA levels or loss of cortical inhibition in the visual system have been associated with visual hallucinations in PD and DLB respectively,^89, 90^ while postmortem studies in DLB report downregulation of postsynaptic GABAergic markers despite preserved interneuron density.^70^ These findings point to finely tuned inhibitory circuitry as a specific vulnerability target in DLB. Clinical sensitivity of DLB patients to GABAergic agents such as benzodiazepines, which might lead to increased sedation and worsened cognitive symptoms,^91, 92^ further underscores the fragility of this system. On the other hand, given the association between GABA and clinical features such as somnolence, another hypothesis might be that the observed association with GABAergic markers may reflect phenotypic expression influencing clinical diagnosis rather than underlying disease-specific biology. Beyond GABA, the neurotransmitter system association analysis also revealed that regions with cortical thinning in DLB also showed higher expression of D2 receptors. As hypersensitivity to antipsychotic drugs has been well recognized in DLB ^1, 93^ and D2 receptors are their primary target, this spatial overlap may also point to a potential pathophysiological basis for the neuroleptic sensitivity observed in patients with DLB.

Regions with greater cortical thinning in DLB also showed a serotoninergic pattern characterized by higher normative 5-HT_1A_ and 5-HT4 receptor density and lower 5-HT_1B_ receptor density. This is consistent with pathological models of misfolded alpha-synuclein distribution showing early involvement of the raphe nuclei,^94^ which may disrupt serotoninergic transmission and contribute to depressive symptoms in Lewy body disease.^95^ It also aligns with reports of increased 5-HT_1A_ receptor density in temporal regions in DLB and PD dementia with depressive symptoms.^96^ The association with 5-HT4 is also notable because 5-HT4 agonists have been explored in clinical trials due to their potential to stimulate acetylcholine release, with memory-enhancing effects thought to be mediated through medial temporal lobe structures,^97^ although this trial was terminated. The specificity of these serotoninergic association may reflect their regional distribution, with 5-HT_1A_ and 5-HT4 being preferentially expressed in temporal cortical areas compared to other cortical regions and overlapping with DLB cortical thinning. 5-HT_1B_ is comparatively less expressed in these same regions. By contrast, 5-HT_2A_, whose activation is associated with hallucinatory effects, and which has been implicated in hallucinations in Lewy body diseases,^98^ is highly expressed across both the temporal and frontal cortices^99^ and was therefore not significantly associated with cortical thinning in our analysis. Thus, the association between normative serotoninergic architecture and DLB-related cortical thinning appears to depend on receptor distributions with stronger specificity for temporal cortical regions.

Further supporting the biological specificity of our findings, nearly half of the DLB-enriched genes were significantly overexpressed in brain tissue relative to peripheral organs and were enriched for the regulation of trans-synaptic signalling. These transcriptomic findings align with the prion-like spread of misfolded proteins in neurodegenerative disorders, which relies on the idea that proteins may spread trans-synaptically along connected networks. That atrophy in DLB may follow such a prion-like constraint was substantiated by our connectivity analysis, which demonstrated that regional cortical thinning in DLB did significantly correlate with the degree of thinning in structurally connected neighbours. By contrast, no significant association was observed after spatial autocorrelation correction for the functional connectivity-weighted connectome, suggesting that anatomical connectivity may more strongly constrain the spatial distribution of cortical thinning in DLB. However, the functional connectivity association remained close to significance after spatial correction (P_uncorr_ = 0.008, P_spatial_ = 0.071), indicating that functional network organization may still contribute to DLB-related atrophy patterns. Taken together, these findings align with growing evidence from alpha-synuclein seeding models, which show that pathological aggregates propagate along axonal projections in a connectivity-dependent manner.^13-16, 20, 22, 100-102^ In iRBD, several brain changes have been reported as predictive of DLB development.^29, 102-104^ In silico models combining structural connectivity and gene expression have also demonstrated that the similarity between alpha-synuclein spread and observed atrophy patterns predicts cognitive decline,^20^ a trajectory more strongly associated with conversion to DLB than PD.^55, 105^ Taken together, these converging lines of evidence suggest that the selective vulnerability observed in DLB arises not from isolated regional pathology but from an interplay between molecular predisposition and the architecture of large-scale brain networks. This integrated model of disease opens the door to predictive frameworks for tracking brain disease evolution and suggests that circuit-level therapeutic interventions may hold potential for slowing neurodegeneration in DLB.

This study has some limitations. First, our analyses relied solely on T1-weighted MRI, which captures macrostructural atrophy but no other pathological hallmarks such as neuroinflammation, gliosis, or iron accumulation. Future studies should integrate complementary modalities such as diffusion MRI and quantitative susceptibility mapping to obtain a more comprehensive characterization of neurodegeneration in DLB. Second, underlying co-pathologies, such as AD pathology, may influence cortical atrophy and its biological underpinnings in DLB. In our cohorts, biomarkers for AD pathology were only available for a subset of patients and future studies should address whether the presence of AD co-pathology in DLB influence the biological factors that underlie regional vulnerability. Third, the MRI data were aggregated across multiple cohorts, each with partially differing clinical assessments. All cohorts captured core clinical features of DLB, but this heterogeneity limited our ability to perform unified brain-behaviour correlations. Fourth, transcriptomic associations were derived from healthy donors in the Allen Human Brain Atlas, as our objective was to identify normative molecular features conferring vulnerability. Although this approach is conceptually appropriate for investigating predisposition, future postmortem studies in DLB can help to determine whether these normative signatures are preserved or are altered in the diseased brain. However, the limited spatial coverage of postmortem data in DLB currently precludes the kind of whole-brain modelling used in this study. Fifth, our analyses focused on cortical brain changes because cortical Lewy body pathology is a defining feature of DLB.^106^ Moreover, previous MRI-derived subtypes of brain atrophy progression have shown that iRBD patients evolving toward an early dementia-first phenotype exhibit extensive cortical changes associated with cognitive decline before subsequent involvement of subcortical structures, where parkinsonian motor features emerge and contribute to a DLB diagnosis.^55^ This provides insight into the potential determinants of cortical brain changes in DLB, but future studies should further investigate whether similar vulnerability profiles and prion-like mechanisms also shape changes in subcortical, brainstem, and white matter structures affected in DLB. Sixth, our connectomic analyses relied on normative rather than patient-specific structural and functional connectivity data, as done previously.^20, 21, 59^ This allowed testing whether canonical connectivity patterns aligned with cortical thickness in DLB but did not capture individual connectivity disruptions that may shape patient-specific thinning patterns. Currently, generating reliable connectomes in DLB is a challenging task because substantial atrophy can affect segmentation, registration, tractography, and connectivity estimation. Future studies should investigate how deep learning-based processing approaches may support better derivation of such information in the DLB brain.

## Conclusions

In summary, our study reveals that cortical thinning in DLB follows a distinct pattern shaped by specific gene expression, neurochemical, and anatomical features. We found that vulnerable regions are enriched for mitochondrial and synaptic genes, overlap with AD and PD pathways, and are embedded in the structural connectome. We also identified GABAergic genes preferentially expressed in brain tissue that may reflect DLB-specific molecular vulnerability. Together, these findings highlight how intrinsic molecular and network architecture contribute to selective neurodegeneration in DLB, offering new avenues for refined prognostic and therapeutic strategies.

## List of abbreviations

AD: Alzheimer’s disease
DLB: dementia with Lewy bodies
MRI: magnetic resonance imaging
PD: Parkinson’s disease

## Declarations

### Ethics approval and consent to participate

All participants provided written informed consent in accordance with institutional ethics protocols. The original study protocols received approval from local research ethics committees, and the present study was approved by the Research Ethics Boards of the McGill University Health Centre and the Quebec Integrated University Centre for Health and Social Services of Northern Island of Montreal.

### Consent for publication

Not applicable

### Authors’ contributions

A.De. and S.R. wrote the main manuscript text. A.De., S.J., C.T., A.V., M.F. and S.R. performed the processing and data analysis. J.P.T., J.T.O., M.F., A.T., P.C.D., R.C., H.C., A.D., and R.B.P. performed data acquisition and interpretation. S.R. supervised the project and funding.

## Acknowledgments

Outside the submitted work, Stephen Joza received support for attending meetings and/or travel from the American Academy of Neurology and Parkinson’s Canada and was supported by an Edmont J. Safra Fellowship in Movement Disorders from the Michael J. Fox Foundation. John-Paul Taylor has acted as the UK Chief Investigator for ReWIND-DLB, CervoMed, acted as a consultant for Eisai and received speaker fees from GE Healthcare and Bial. John T. O’Brien received consulting fees from Biogen and acted as a consultant for Roche, GE Healthcare, and Okwin. He received honoraria for lectures from GE Healthcare, serves on advisory boards or DSMBs for TauRx and Novo Nordisk, chairs the Research Strategy Council of the UK Alzheimer’s Society, and received research support from Avid/Lilly, Merck, UCB, and Alliance Medical. Paul C. Donaghy received grants or contracts, paid to his institution, from Alzheimer’s Research UK, the Michael J. Fox Foundation, the Alzheimer’s Society, and GE Healthcare, and received an honorarium for a lecture at the Lewy Body Masterclass (paid to his institution). Richard Camicioli serves (unpaid) on the Research and Scientific Advisory Board of Parkinson Canada. Howard Chertkow is the Scientific Director of CCNA (unpaid) and principal investigator or co-investigator on major research grants including from CIHR ($20.3M, 2024–29), Alzheimer’s Society of Canada, BrightFocus, and NIH. He has also received funding for multi-site clinical trials sponsored by IntelGenX, Alector, Eli Lilly, Biogen, Hoffman LaRoche, and Anavex. He serves on advisory boards for Lilly and Eisai (personal payment). Alain Dagher received travel support from the Michael J. Fox Foundation. Ronald B. Postuma received grants from the CIHR, Michael J. Fox Foundation, NIH, Roche Diagnostics, and the Weston Foundation. He received consulting fees from Novartis, Eisai, Merck, Vaxxinity, BMS, Ventus, Korro, Vanqua, Roche, Regeneron, Helicon, Epic, and Clinilabs. He holds leadership roles with Parkinson Canada, the Michael J. Fox Foundation, MDS, Movement Disorders journal, and the International REM Sleep Behaviour Disorder Study Group (IRBDSG). Shady Rahayel holds a Junior 1 research scholar award from the Fonds de recherche du Québec – Santé (FRQS). Aline Delva, Christina Tremblay, Andrew Vo, Marie Filiatrault, Michael Firband and Alan Thomas report no conflicts of interest.

## Acknowledgements and Funding Sources

The authors thank all participants for their contribution. We acknowledge the donation in memoriam of Gaetan Boulianne and Pierre-Claude Durivage to the Foundation of Hopital du Sacre-Coeur de Montreal, in support of PD research at the Center for Advanced Research in Sleep Medicine.

